# Reference values and determinants of fractional exhaled nitric oxide in a representative adult population in Western Sweden

**DOI:** 10.1101/2024.11.04.24316695

**Authors:** Reshed Abohalaka, Selin Ercan, Lauri Lehtimäki, Saliha Selin Özuygur Ermis, Daniil Lisik, Muwada Bashir Awad Bashir, Radhika Jadhav, Linda Ekerljung, Göran Wennergren, Jan Lötvall, Teet Pullerits, Helena Backman, Madeleine Rådinger, Bright Ibeabughichi Nwaru, Hannu Kankaanranta

## Abstract

**Background:** Fractional exhaled nitric oxide (FeNO) is used to differentiate asthma inflammatory phenotypes and guide its management. However, data on FeNO reference values in a representative adult population is limited.

**Objective:** To derive reference values and determinants of FeNO in a representative adult population.

**Methods:** The West Sweden Asthma Study is a clinical-epidemiological population- representative study of randomly selected adults in Western Sweden. From this cohort, 943 subjects participated in comprehensive clinical investigations, including skin prick testing (SPT), specific immunoglobulin E (sIgE) analysis, and FeNO measurement. Clinical allergy was defined as co-occurrence of atopy (positivity to SPT or sIgE) and self-reported allergic symptoms to the same allergen family. FeNO levels were analysed in relation to the presence or absence of clinical allergy, asthma, and other factors.

**Results:** The 95^th^ percentile of FeNO ranged from 34 to 52 parts per billion (ppb) in the entire sample (*N*=943), and from 26 to 37 ppb among individuals without clinical allergy, asthma, or chronic obstructive pulmonary disease (COPD) (*n*=587), depending on age. Sex, smoking, clinical allergy, atopy, asthma, and hypertension influenced FeNO levels, meanwhile, age, asthma, clinical allergy, and reversibility- related variables were significant determinants of FeNO levels.

**Conclusion:** The 95^th^ percentile (upper normal limit) for FeNO ranges from 34 to 52 ppb overall, and from 26 to 37 ppb in those without clinical allergy, asthma, or COPD, depending on age. These findings provide a guide for interpreting FeNO in the general population and in asthma and COPD clinics.

## Introduction

Nitric oxide is a cellular signalling molecule that plays a pivotal role in the physiological regulation of respiratory tract function and inflammatory processes [1,2]. Measurement of fractional exhaled nitric oxide (FeNO) correlates with induced- sputum eosinophilia [3] as well as with airway hyperresponsiveness [3,4]. Furthermore, the measurements are characterized by ease of execution and high patient acceptance [5]. FeNO thus stands as a promising biomarker differentiating asthma from other respiratory diseases [6,7]. While using FeNO to confirm an asthma diagnosis remains a subject of controversy [2], it is acknowledged as a valuable biomarker for identifying asthma phenotypes and guiding asthma treatment [8,9]. Elevated FeNO levels are associated with poor asthma control [4], as well as reduction in FeNO levels following inhaled steroid treatment [10,11]. Beyond asthma, elevated FeNO levels are seen in chronic obstructive pulmonary disease (COPD) patients [12] and a correlation between FeNO levels and sputum eosinophils in smoking COPD patients has been reported [13]. These observations highlight the potential of FeNO as a marker for Type 2 (T2) airway inflammation [3,14,15].

Many of the few reports that have attempted to establish reference values for FeNO in the general population are not population representative studies [16,17]. Meanwhile, the rest confine their allergy definition exclusively to atopy [18,19], omit upper limits of normal, exclude smokers, or neglect to examine the effects of allergy at all [16,20], thus ignoring clinical variables that may exert a significant influence on FeNO levels. These studies were able to detect an overlap in FeNO values between their “healthy” individuals and those suffering from asthma. Consequently, guidelines recommended the adoption of cut-off values instead of reference values for interpreting FeNO levels. For instance, the American Thoracic Society (ATS) suggests using cut-offs of FeNO >25 and >50 parts per billion (ppb) to determine the response to corticosteroid therapy in adults as indeterminate or likely, respectively [21]. According to the Global Initiative for Asthma (GINA), a FeNO concentration ≥ 20 ppb as part of systematic assessment of difficult-to-treat asthma under regular medication is considered high, along with other biomarkers indicative of T2 immune response [22]. The European Respiratory Society (ERS) recommends a cut-off of 40 ppb for diagnostics [23]. The determination of these cut-off points does not derive from reference values established within the general population [24]. Instead, the selection of these cut-off values is predicated upon achieving an optimal balance in relation to the correlation between FeNO levels and sputum eosinophils, as well as in the context of diagnosing asthma within studies with relatively modest sample sizes [24,25]. Furthermore, various factors influence FeNO values in healthy populations, including age, height, ethnicity, sex, atopy, and smoking [16,18,19,26,27]. Despite this, prevailing clinical guidelines advocate for either age-specific threshold values [24] or a singular cut-off point for interpreting FeNO results [22].

Therefore, this work aimed to ascertain the reference values of FeNO based on a representative adult population in Western Sweden, while considering the impact of various allergic and non-allergic, inflammatory and non-inflammatory factors.

## Methods

### Study area and population

The West Sweden Asthma Study (WSAS) is a population-based cohort study, which has been previously described in detail [28]. Briefly, WSAS follows randomly selected individuals aged 16 to 75 years at baseline. Initiated in 2008, 30,000 subjects within this age group were chosen from the Swedish Population Register to participate in a postal survey. The selection process considered age and gender stratification to ensure a representative sample of the population in Västra Götaland county, comprising nearly a fifth of the total Swedish population.

From the survey responders, a random subset of 2,000 subjects was invited to undergo comprehensive clinical examinations, and 1,172 took part between 2009 and 2012. Ultimately, 943 participants successfully completed all clinical measurements, including the measurement of FeNO (**Figure 1**). All participants signed an informed consent to the study protocol, approved by the regional ethics board in Gothenburg, Sweden.

**Figure 1:**
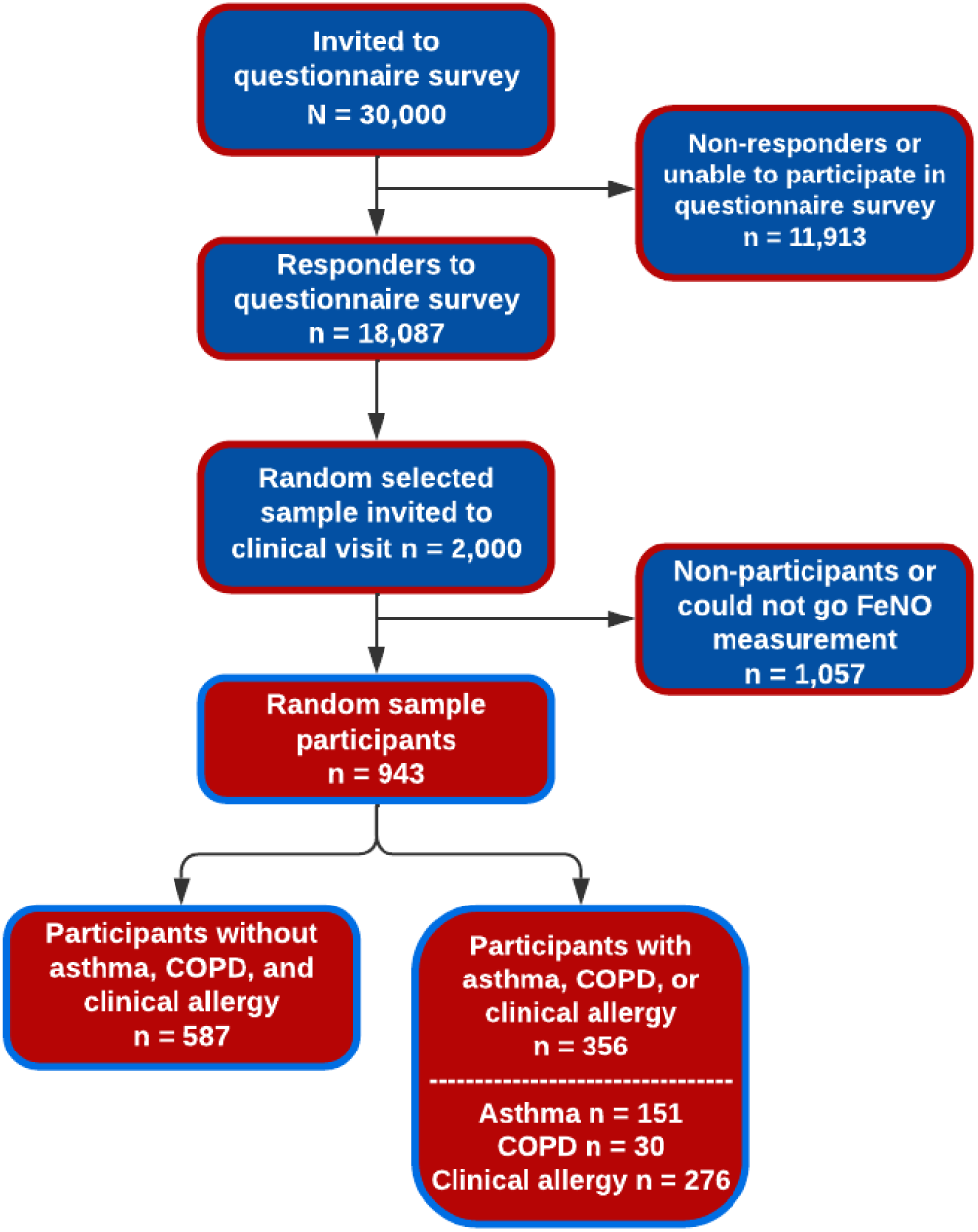
The flowchart illustrating the establishment of participant cohorts emanates from a systematically sampled populace in the Western Sweden Asthma Study (WSAS).

### Clinical examinations

The clinical investigations included haematological examinations for blood differential cell counts, allergic sensitization evaluations through skin-prick tests (SPTs) and specific immunoglobulin E (sIgE) levels, anthropometric assessments involving measurements of height and weight, determination of FeNO, spirometry, and methacholine challenge. In addition, clinical interviews were conducted, along with the administration of questionnaires including, but not limited to, respiratory diseases, allergy, symptoms, and comorbidities.

### Measurement of lung function and FeNO

Spirometry measurements were conducted using a MasterScope spirometer (Jaeger, Höchberg, Germany), adhering to the guidelines stipulated by the ERS/ATS [29,30]. The Global Lung function Initiative’s normal equation for Caucasians was utilized for reference values [31], enabling the calculation of Forced Expiratory Volume in 1 second (FEV_1_) percentage of predicted normal value (FEV_1_% predicted). Reversibility testing was performed 15 minutes thereafter. Methacholine challenge was executed by Spira equipment (Spira Respiratory Care Center Ltd, Hämeenlinna, Finland) adhering to ERS guidelines [32].

FeNO was assessed using a chemiluminescence analyser (NiOX VERO®, Aerocrine, Morrisville, NC, USA). These FeNO measurements were conducted prior to spirometry. The analyser sensor was replaced after 300 measurements or one year, whichever came first. All procedures adhered to the latest ATS guidelines [21,24]. During the measurement, subjects exhaled against a mouth pressure of 5 cm H_2_O aiming at a flow rate of 50 mL/s. NO concentration was measured during the

last 5 seconds of a 10 s period when flow rate was between 45 and 55 ml/s. If the FeNO measurement did not meet the predefined quality criteria [24], the measurement was repeated until an acceptable result was obtained.

### Assessment of sensitization and clinical allergy

Sensitization was assessed through the determination of sIgE levels and/or skin prick tests for 11 aeroallergens (Details are provided in the **supplementary file**). *Clinical allergy* was defined by the presence of allergic sensitization (positive SPT or sIgE to any allergen; atopy), and self-reported allergic symptoms attributable to the same allergen family. These symptoms, evaluated during the clinical interview prior to the sensitization test results, included ocular manifestations, nasal discomfort, various forms of allergic nasal expressions, pruritus in the oral or pharyngeal region, respiratory challenges, exacerbation of asthma symptoms, pruritic skin rash, and disruptions in gastrointestinal function.

### Definitions of diseases

*Asthma* was defined as meeting any of the following criteria: 1) documented diagnosis of asthma by a healthcare professional or a history of asthma accompanied by respiratory symptoms or use of asthma medication in the past 12 months; 2) respiratory symptoms or use of asthma medication in the past 12 months, coupled with a positive reversibility test (12% and 200 ml increase in FEV_1_); or 3) respiratory symptoms or use of asthma medication in the past 12 months, accompanied by a positive methacholine challenge indicative of asthma. *COPD* was defined by a post-bronchodilator FEV_1_/Forced Vital Capacity (FEV_1_/FVC) ratio of less than 0.7, along with a smoking history of 10 or more pack-years [33]. *Metabolic disease* was defined as having obesity (body mass index [BMI] ≥30 kg/m2) or any of the following self-reported conditions: hypertension, hyperlipidaemia, or diabetes [34].

### Statistical analyses

In the establishment of reference parameters, the presumption is that the spectrum of values derived from a cohort deemed healthy serves as a proxy for normative values. Consequently, individuals manifesting values beyond the norm, typically delineated by the central 90% of the range of values in the healthy population (upper limit of normal or 95^th^ percentile and lower limit of normal or 5^th^ percentile), are frequently labelled abnormal [35]. As a result, our reporting includes the 50^th^ 75^th^, and particularly the 95^th^ percentiles of FeNO values, with a specific emphasis on the upper limit of normal, defined as the 95^th^ percentile. Because FeNO values showed a right-skewed distribution, we applied a logarithmic transformation to the dataset, following the method described previously [36]. Percentiles were then calculated from the transformed data for the entire sample and for each 10-year age group. Afterward, the percentile values were converted back to their original scale and used to plot the percentile curves.

Multiple linear regression analyses were conducted using enter method to discern factors associated with higher FeNO values considering potential confounders. Mean comparisons were depicted with corresponding standard deviations and assessed employing independent Student’s t-tests. Statistical significance was determined at *p*<0.05. All statistical analyses were performed using SPSS 29.0 (IBM Corp, New York, USA).

## Results

### Characteristics of the random sample

The random sample, which underwent comprehensive clinical investigation, comprised of 943 individuals (mean age = 50.5 years, SD = 15.5), with 47.7 % (*n* = 450) being males. More than half of the participants (51.6 %, *n* = 487) reported no history of smoking, while 10.8% (*n* = 102) were current smokers. Individuals presenting with clinical allergy tended to be younger and predominantly male, with a lower incidence of smoking history compared to those without clinical allergy. Moreover, the prevalence of asthma was found to be elevated among participants exhibiting clinical allergy compared to those without clinical allergy. For further insights into lung function, allergic parameters, and comorbidities among participants with or without clinical allergy, see **Table 1**.

**Table 1.**
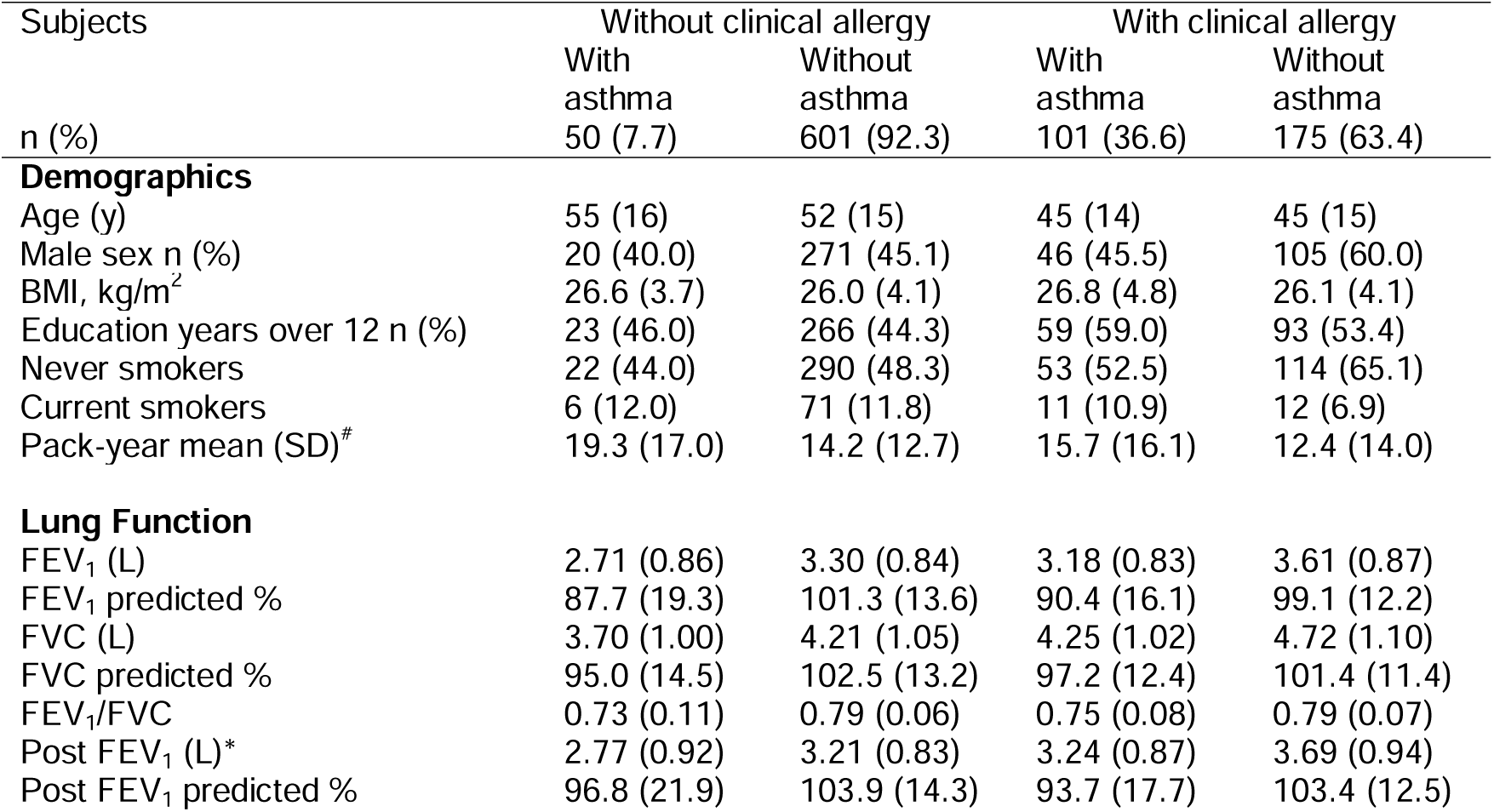
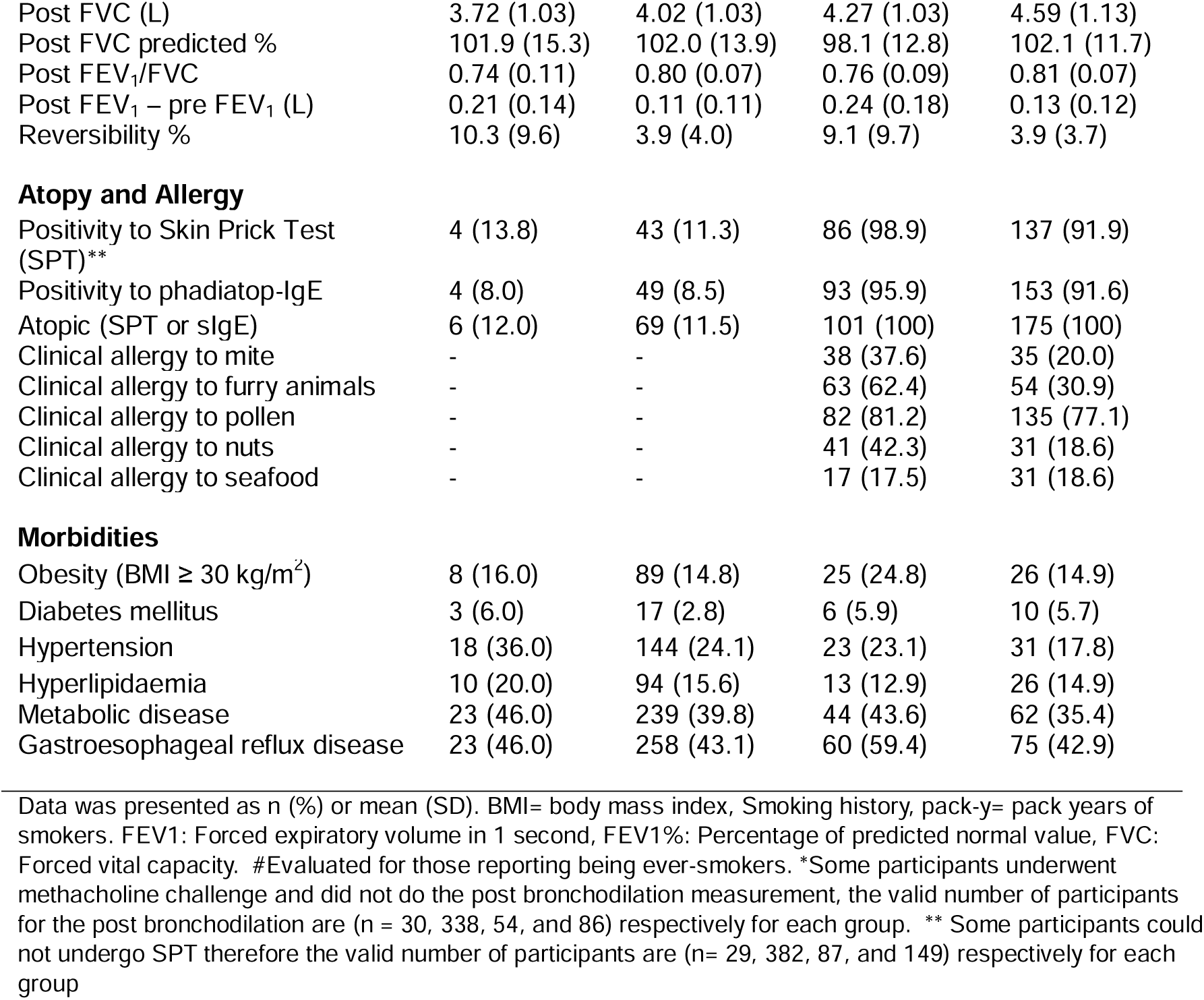
Characteristics of WSAS I random sample subjects with or without asthma and clinical allergy (N=943).

| Subjects | Without clinical allergy |  | With clinical allergy |  |
| --- | --- | --- | --- | --- |
|  | With<br>asthma | Without<br>asthma | With<br>asthma | Without<br>asthma |
| $n$ (%) | 50 (7.7) | 601 (92.3) | 101 (36.6) | 175 (63.4) |
| **Demographics** |  |  |  |  |
| Age (y) | 55 (16) | 52 (15) | 45 (14) | 45 (15) |
| Male sex $n$ (%) | 20 (40.0) | 271 (45.1) | 46 (45.5) | 105 (60.0) |
| BMI, kg/m<sup>2</sup> | 26.6 (3.7) | 26.0 (4.1) | 26.8 (4.8) | 26.1 (4.1) |
| Education years over 12 $n$ (%) | 23 (46.0) | 266 (44.3) | 59 (59.0) | 93 (53.4) |
| Never smokers | 22 (44.0) | 290 (48.3) | 53 (52.5) | 114 (65.1) |
| Current smokers | 6 (12.0) | 71 (11.8) | 11 (10.9) | 12 (6.9) |
| Pack-year mean (SD)* | 19.3 (17.0) | 14.2 (12.7) | 15.7 (16.1) | 12.4 (14.0) |
| **Lung Function** |  |  |  |  |
| FEV<sub>1</sub> (L) | 2.71 (0.86) | 3.30 (0.84) | 3.18 (0.83) | 3.61 (0.87) |
| FEV<sub>1</sub> predicted % | 87.7 (19.3) | 101.3 (13.6) | 90.4 (16.1) | 99.1 (12.2) |
| FVC (L) | 3.70 (1.00) | 4.21 (1.05) | 4.25 (1.02) | 4.72 (1.10) |
| FVC predicted % | 95.0 (14.5) | 102.5 (13.2) | 97.2 (12.4) | 101.4 (11.4) |
| FEV<sub>1</sub>/FVC | 0.73 (0.11) | 0.79 (0.06) | 0.75 (0.08) | 0.79 (0.07) |
| Post FEV<sub>1</sub> (L)* | 2.77 (0.92) | 3.21 (0.83) | 3.24 (0.87) | 3.69 (0.94) |
| Post FEV<sub>1</sub> predicted % | 96.8 (21.9) | 103.9 (14.3) | 93.7 (17.7) | 103.4 (12.5) |
| Post FVC (L) | 3.72 (1.03) | 4.02 (1.03) | 4.27 (1.03) | 4.59 (1.13) |
| Post FVC predicted % | 101.9 (15.3) | 102.0 (13.9) | 98.1 (12.8) | 102.1 (11.7) |
| Post FEV<sub>1</sub>/FVC | 0.74 (0.11) | 0.80 (0.07) | 0.76 (0.09) | 0.81 (0.07) |
| Post FEV<sub>1</sub> – pre FEV<sub>1</sub> (L) | 0.21 (0.14) | 0.11 (0.11) | 0.24 (0.18) | 0.13 (0.12) |
| Reversibility % | 10.3 (9.6) | 3.9 (4.0) | 9.1 (9.7) | 3.9 (3.7) |
| Atopy and Allergy |  |  |  |  |
| Positivity to Skin Prick Test (SPT)** | 4 (13.8) | 43 (11.3) | 86 (98.9) | 137 (91.9) |
| Positivity to phadiatop-IgE | 4 (8.0) | 49 (8.5) | 93 (95.9) | 153 (91.6) |
| Atopic (SPT or sIgE) | 6 (12.0) | 69 (11.5) | 101 (100) | 175 (100) |
| Clinical allergy to mite | - | - | 38 (37.6) | 35 (20.0) |
| Clinical allergy to furry animals | - | - | 63 (62.4) | 54 (30.9) |
| Clinical allergy to pollen | - | - | 82 (81.2) | 135 (77.1) |
| Clinical allergy to nuts | - | - | 41 (42.3) | 31 (18.6) |
| Clinical allergy to seafood | - | - | 17 (17.5) | 31 (18.6) |
| Morbidities |  |  |  |  |
| Obesity (BMI ≥ 30 kg/m<sup>2</sup>) | 8 (16.0) | 89 (14.8) | 25 (24.8) | 26 (14.9) |
| Diabetes mellitus | 3 (6.0) | 17 (2.8) | 6 (5.9) | 10 (5.7) |
| Hypertension | 18 (36.0) | 144 (24.1) | 23 (23.1) | 31 (17.8) |
| Hyperlipidaemia | 10 (20.0) | 94 (15.6) | 13 (12.9) | 26 (14.9) |
| Metabolic disease | 23 (46.0) | 239 (39.8) | 44 (43.6) | 62 (35.4) |
| Gastroesophageal reflux disease | 23 (46.0) | 258 (43.1) | 60 (59.4) | 75 (42.9) |

Data was presented as n (%) or mean (SD). BMI= body mass index, Smoking history, pack-y= pack years of smokers. FEV<sub>1</sub>: Forced expiratory volume in 1 second, FEV<sub>1</sub> %: Percentage of predicted normal value, FVC: Forced vital capacity. #Evaluated for those reporting being ever-smokers. \*Some participants underwent methacholine challenge and did not do the post bronchodilation measurement, the valid number of participants for the post bronchodilation are (n = 30, 338, 54, and 86) respectively for each group. \*\* Some participants could not undergo SPT therefore the valid number of participants are (n= 29, 382, 87, and 149) respectively for each group

### Fractional exhaled nitric oxide in the random sample

The distribution of FeNO levels within the random sample was right-skewed, as depicted in **Figure 2**. Consequently, percentiles were calculated from the dataset after logarithmic transformation, with a median value of 16.9 ppb. The lower and upper normal limits corresponding to the 5^th^ and 95^th^ percentiles were determined to be 7.0 ppb and 41.2 ppb, respectively.

**Figure 2:**
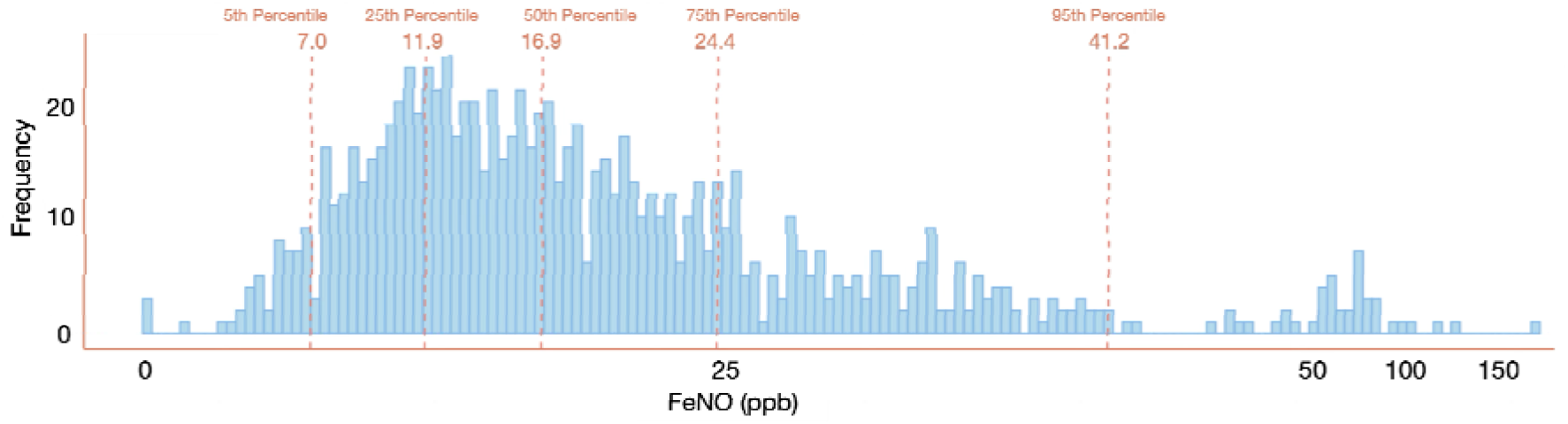
Frequency distribution of Fractional exhaled nitric oxide (FeNO) in the entire random sample. The dotted lines indicate the 5^th^, 25^th^, 50^th^, 75^th^ and 95^th^ percentiles.

The upper normal limit of FeNO within the entire sample exhibited a gradual increase with increasing age after 30 years old, meanwhile, it decreased from 52.2 ppb for 18- 30 years age group to 34.0 ppb for 30-40 years age group (**Figure 3A**). To ascertain the influence of asthma and COPD on FeNO levels, individuals diagnosed with either condition were excluded from the analysis (**Figure 3B**). Remarkably, upper normal limit of FeNO showed quite similar pattern as in the whole population. Following this, we aimed to explore the impact of clinical allergy on FeNO levels by additionally excluding these with clinical allergy (**Figure 3C**). The upper normal limit reduced among individuals without clinical allergy, asthma, and COPD, particularly within the 18-30 years age group, and was highest (37.0) among the 60+ years age group. Lastly, participants lacking atopy had a similar FeNO profile as those without clinical allergy (**Figure 3D**).

**Figure 3:**
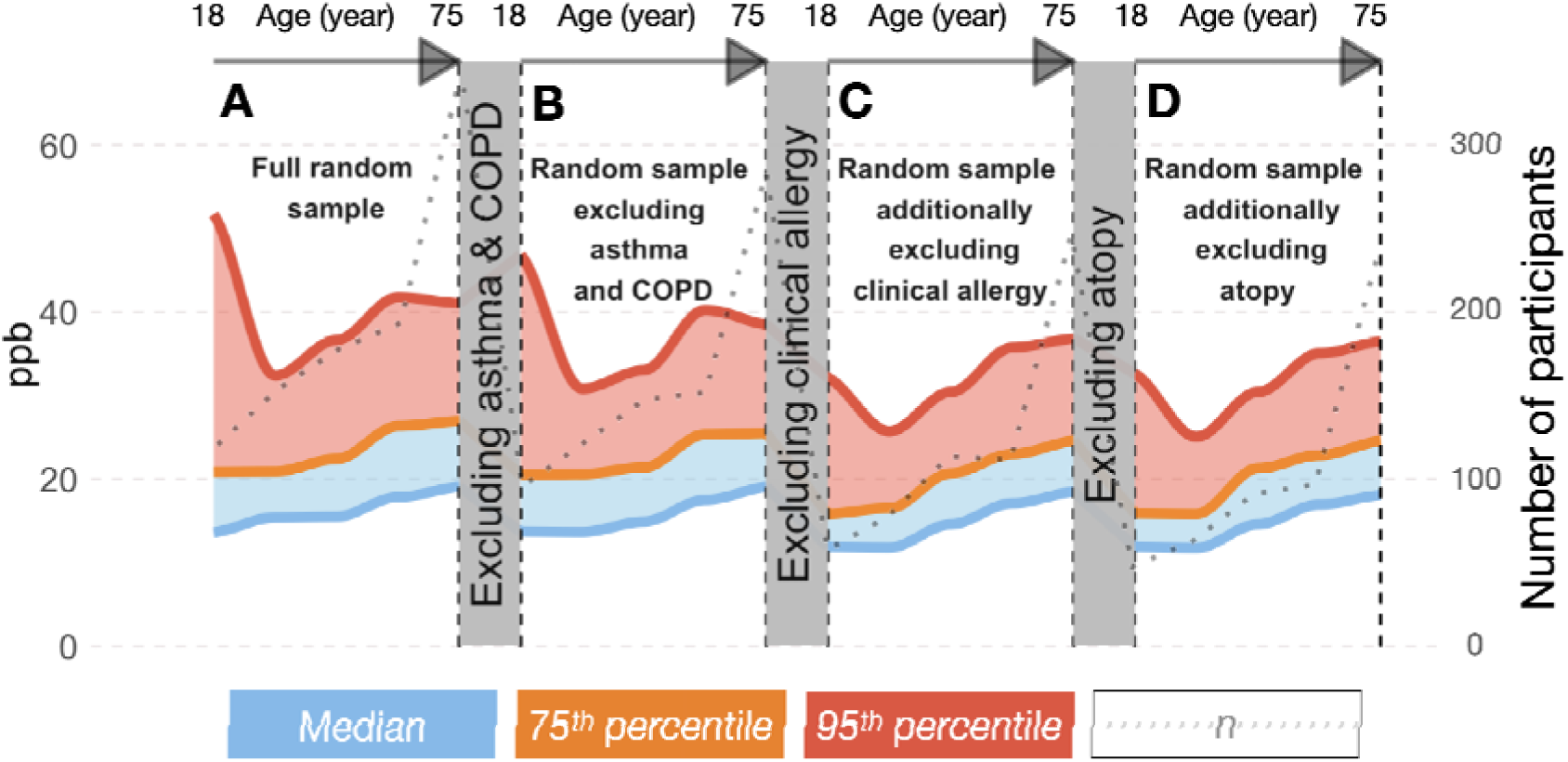
The characteristics of FeNO across different age groups, delineated for (A) the entire random sample (*N*=943), (B) random sample participants excluding asthma and COPD patients (*n*=773), (C) individuals additionally devoid of clinical allergy (*n*=587), and (D) individuals additionally devoid of atopy (*n*=519).

The 75^th^ percentile and the median of FeNO showed similar patterns of change to the 95^th^ percentile with age and for these without asthma, COPD, or clinical allergy in our cohort. However, unlike the 75^th^ percentile and the median, the upper normal limit decreased more remarkably in individuals without clinical allergy, asthma, and COPD, especially those aged 18 to 30. Therefore, we selected individuals without clinical allergy, asthma, and COPD as the “healthy” cohort for our study. For FeNO reference values in different subgroups of our cohort, see **Figure S1.**

### Factors affecting fractional exhaled nitric oxide in the random sample

A comprehensive understanding of the determinants affecting the upper normal limit of FeNO within the general population necessitates a comparative investigation of FeNO upper 95^th^ percentile values across diverse demographic and clinical parameters. However, the sample size in our study was insufficient to draw statistically significant conclusions from such comparisons. However, when comparing the upper 95^th^ percentile values of clinical parameters with adequate sample sizes, the mean showed a similar pattern (**Figure S1B**). Moreover, the upper 95^th^ percentile is a collective outcome and not individually available for each patient. Hence, we opted to conduct a comparative analysis of FeNO mean values to ascertain the directional impact of various demographic and clinical parameters previously identified to influence FeNO levels. Our investigation revealed notable disparities in FeNO levels across different subgroups. Specifically, FeNO levels were significantly higher in males than in females. This difference persisted even when comparing participants of both sexes within the same age and height categories. FeNO levels were significantly higher among both ever and current smokers than among never and non-current smokers. Additionally, FeNO levels were significantly higher among individuals diagnosed with clinical allergy and those with atopy than among those without such conditions, as well as among individuals with asthma than individuals without asthma. Furthermore, our results showed that FeNO levels were elevated in these with hypertension than those without (**Table 2**).

**Table 2.**
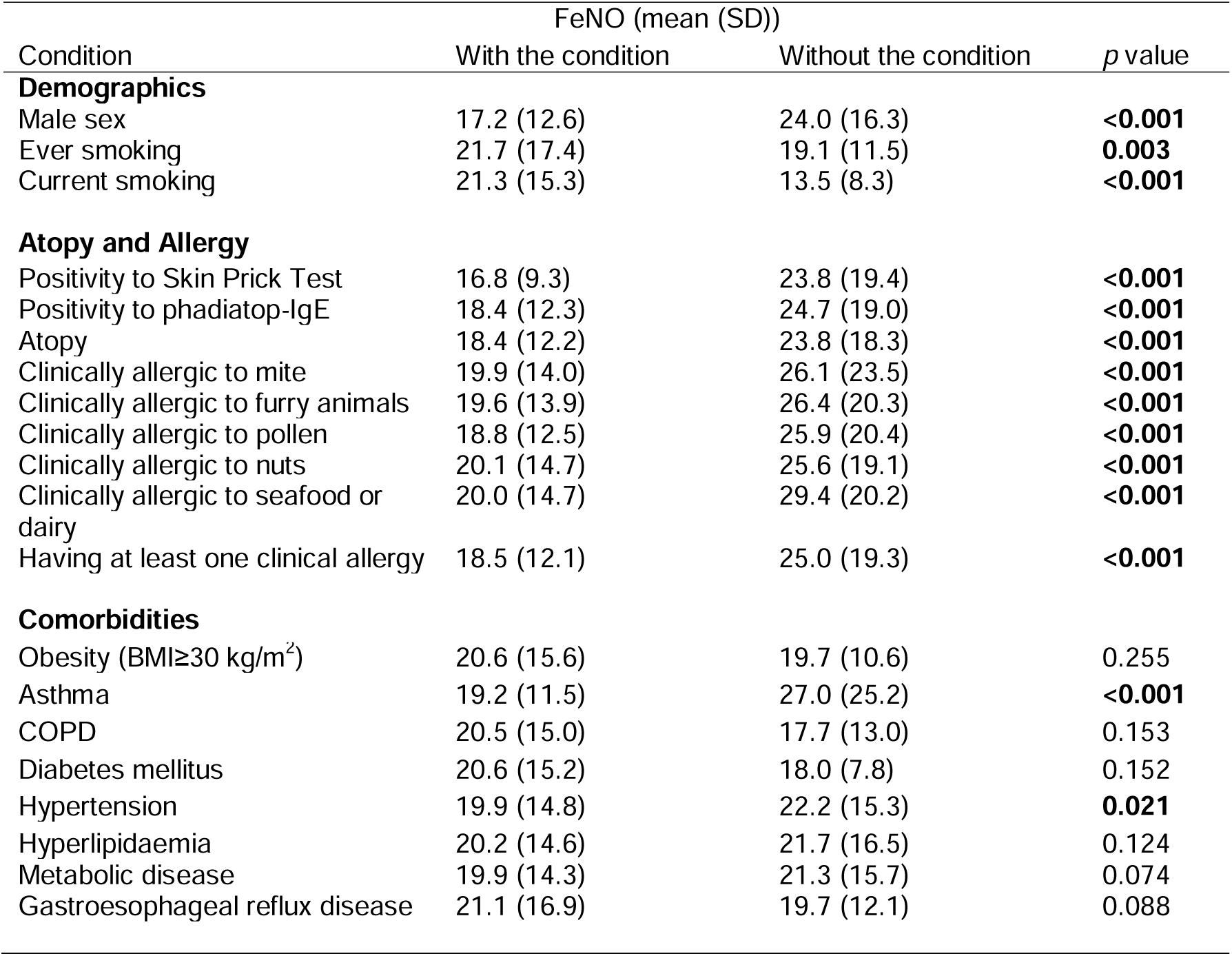
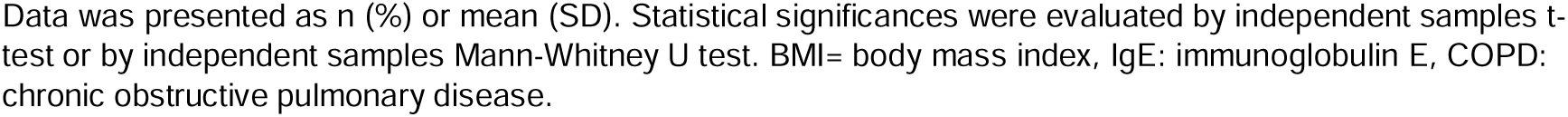
Mean FeNO levels of participants in WSAS random sample according to demographical and clinical features (*n*=943).

### Multiple linear regression model for determinants of FeNO levels

Multiple linear regression analyses were conducted to ascertain the factors associated with increases in FeNO values within the general population, while considering potential covariates based on previous knowledge. The findings indicated that age, asthma, and clinical allergy – although not atopy – emerged as significant determinants of FeNO levels across the entire sample (**Table 3**). Additionally, lung function parameters, such as the percentage of post-FEV_1_ of predicted, the disparity between post- and pre-FEV_1_ in litres, and the percentage of reversibility post bronchodilation test, were identified as significant determinants of FeNO levels across the entire sample.

**Table 3:** Determinants of FeNO levels in multiple linear regression analysis.

|  | Coefficient<br>B | Std. Error | Lower 95%<br>CI | Upper 95%<br>CI | p-value |
| --- | --- | --- | --- | --- | --- |
| **Demographics** |  |  |  |  |  |
| Age | 0.937 | 0.371 | 0.207 | 1.666 | **0.012** |
| Male sex | -0.029 | 3.337 | -6.584 | 6.526 | 0.993 |
| Height | 0.229 | 0.431 | -0.618 | 1.076 | 0.595 |
| Weight | -0.261 | 0.384 | -1.016 | 0.494 | 0.497 |
| BMI | 0.622 | 1.139 | -1.616 | 2.860 | 0.585 |
| Ever smoking | -1.736 | 1.320 | -4.329 | 0.858 | 0.189 |
| Current smoking | -3.231 | 2.069 | -7.295 | 0.834 | 0.119 |
| **Comorbidities** |  |  |  |  |  |
| Asthma | 4.857 | 1.714 | 1.490 | 8.225 | **0.005** |
| COPD | -3.877 | 4.859 | -13.420 | 5.667 | 0.425 |
| Hypertension | 0.162 | 1.722 | -3.220 | 3.544 | 0.925 |
| Diabetes mellitus | -6.529 | 3.611 | -13.622 | 0.565 | 0.071 |
| Atopy | 0.088 | 3.712 | -7.204 | 7.381 | 0.981 |
| Clinical allergy | 5.607 | 2.448 | 0.798 | 10.416 | **0.022** |
| **Biomarkers** |  |  |  |  |  |
| Skin prick test positivity | -1.139 | 3.237 | -7.498 | 5.220 | 0.725 |
| Phadiatop-IgE positivity | 2.786 | 2.547 | -2.216 | 7.788 | 0.274 |
| **Lung function\*** |  |  |  |  |  |
| FEV<sub>1</sub> pred % | 1.976 | 1.384 | -0.743 | 4.695 | 0.154 |
| FVC | 25.717 | 22.169 | -17.827 | 69.262 | 0.247 |
| FVC pred % | 0.558 | 1.446 | -2.282 | 3.398 | 0.700 |
| FEV<sub>1</sub>/FVC | 207.493 | 149.752 | -86.659 | 501.644 | 0.166 |
| Post FEV<sub>1</sub> | 10.473 | 11.040 | -11.212 | 32.158 | 0.343 |
| Post FEV<sub>1</sub> pred % | -3.232 | 1.597 | -6.370 | -0.094 | **0.044** |
| Post FVC | -28.803 | 22.808 | -73.604 | 15.998 | 0.207 |
| Post FVC pred % | 0.585 | 1.518 | -2.397 | 3.567 | 0.700 |
| Post FEV<sub>1</sub>/FVC | -58.906 | 158.921 | -371.067 | 253.255 | 0.711 |
| Post FEV<sub>1</sub>/FVC pred % | 65.375 | 170.610 | -269.747 | 400.497 | 0.702 |
| Post-pre FEV<sub>1</sub> | 60.837 | 27.086 | 7.633 | 114.041 | **0.025** |
| Reversibility % | 1.856 | 0.736 | 0.410 | 3.303 | **0.012** |

**R: 0.489, R Square: 0.240, Adjusted R Square: 0.203**
Variables linked with the elevation of FeNO were examined through linear regression analyses encompassing the entire randomized sample (n = 943). FEV<sub>1</sub>: Forced expiratory volume in 1 second, FEV<sub>1</sub>%: Percentage of predicted normal value, FVC: Forced vital capacity. \*Pre FEV<sub>1</sub> and pre FEV<sub>1</sub>/FVC predicted variables were excluded from the regression analysis due to multicollinearity reasons. Data are presented as the unstandardized b coefficient, standard error of b, and 95% CI.

In addition, we explored the determinants of FeNO levels excluding individuals with asthma, COPD, and allergies. Our analysis using multiple linear regression demonstrated that, in the absence of asthma, allergies or COPD, nearly all variables listed in **Table 3** were significant determinants of FeNO levels (**Table S1**). Within this subgroup, FeNO levels were found to be influenced by both age and height, increasing with the elevation of either parameter (**Figure 4**).

**Figure 4:**
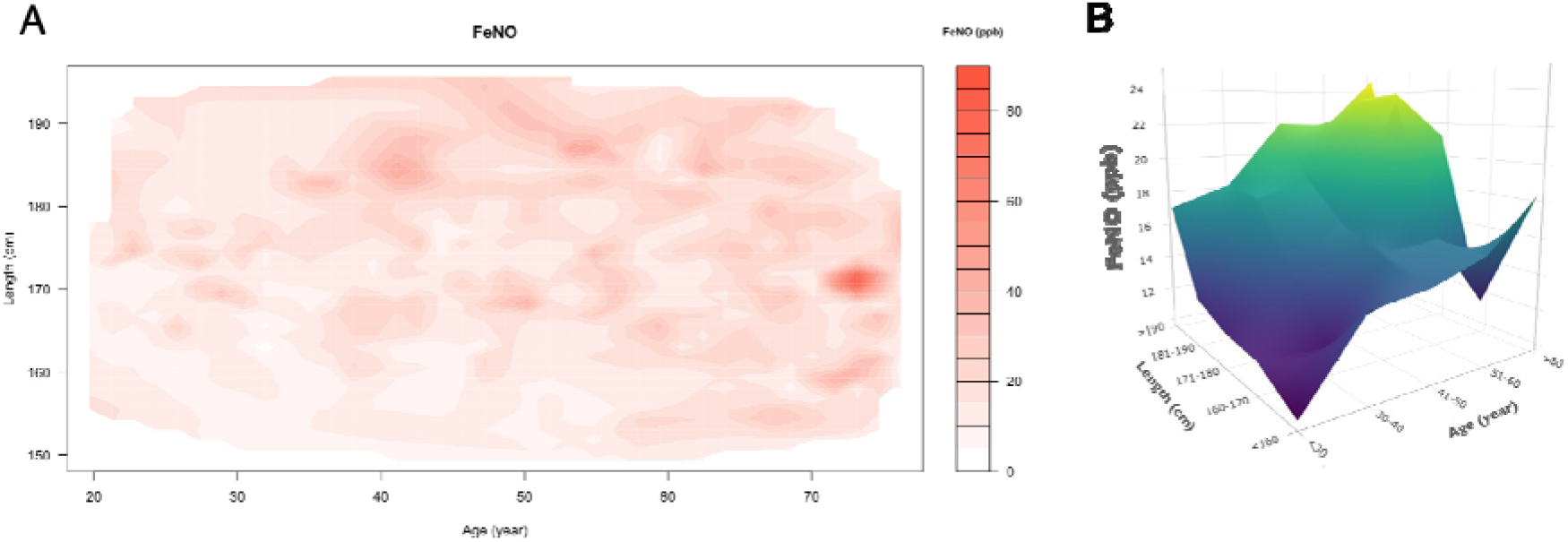
The distribution of FeNO among participants categorized by age and height, excluding those participants with asthma, COPD, and clinical allergy. The distribution is presented in two formats: (A) continuous variables of age and height represented through a heatmap displaying FeNO levels, and (B) age and height grouped variables through a surface expressing the mean value of FeNO.

## Discussion

In our study, the 95^th^ percentile of FeNO ranged from 34 to 52 ppb in the entire sample and from 26 to 37 ppb among individuals without clinical allergy, asthma, and COPD, depending on age. Sex, smoking, clinical allergy, atopy, asthma, and hypertension influenced FeNO levels in the total random sample. Additionally, age, asthma, clinical allergy, the percentage of predicted post-FEV_1_, and reversibility in the bronchodilatation test were significant determinants of FeNO levels across the full sample. These results provide reference values and patterns of FeNO levels in the general population considering various background factors and relevant conditions/comorbidities. This is crucial since FeNO is a key potential biomarker for T2-airway inflammation in asthma and COPD patients [7,13–15].

Few studies have attempted to establish reference values for FeNO among adults. The upper limit of FeNO in non-smoking individuals without allergies ranged from 37.4 to 47.4 ppb depending on gender [19], and from 38.3 to 62 ppb depending on height and gender [37]. In more stringent criteria, where participants were free from infections and medication usage as well, the upper limit was 19.7 ppb [38]. However, there are disparities in participant numbers and the scope of statistical variables considered across these studies. Furthermore, the sample selection process in these studies is not optimal, rendering them non-representative of the general population and limiting their value [24].

Olin et al. [20] conducted a study aimed at formulating an equation to ascertain the upper limit of normal FeNO levels within a cohort devoid of atopic individuals, thus representing a “healthy” population. Their investigation revealed a notable variability in the anticipated upper limit of normal FeNO levels, ranging from 24 ppb to 54 ppb, contingent upon the age and height of the participants. Conversely, Brody et al. [16] also documented that within the adult population, the upper limit of normal FeNO levels, identified using the 97.5^th^ percentile, spanned from 39.5 to 47.7 ppb, with variations correlated with age. These ranges notably exceed the reference value documented in our study and could potentially influence the ATS to advise against the utilization of reference values in interpreting FeNO levels, as they overlap with those calculated from participants with a history of asthma [24]. This discrepancy could potentially arise from methodological differences. Notably, Olin et al. and Brody et al. excluded individuals with a history of smoking, a factor known to reduce FeNO levels, which could potentially skew the variability in FeNO levels, rendering it unrepresentative of the general population. The inclusion of smokers in the determination of FeNO reference values is essential, since smoking is associated with the T2-low asthma phenotype [39]. Moreover, Brody et al. employed exclusion criteria that solely encompassed participants with respiratory conditions and symptoms, while allergic or atopic individuals, who may exhibit elevated FeNO levels, were not excluded. Conversely, our study aimed to establish the upper limit of normal FeNO levels in a healthy population defined by more practical criteria as the absence of asthma, COPD, and clinical allergy, resulting in a range of 26 ppb to 37 ppb, depending on age.

Although age, height and sex are widely reported as influential factors on FeNO levels [16,19,40], Olin et al. [20] found no discernible differences between males and females of similar age and height. Conversely, Travers et al. [19] and Taylor et al. [40] consistently reported elevated FeNO levels in males. Our study corroborates these findings, revealing significantly higher FeNO levels among males within the entire random sample, even when comparing participants of both sexes within identical age and height categories. Moreover, the significance remained after adjusting FeNO levels between males and females for height using a generalized linear model. This underscores the potentially clinically significant impact of sex on FeNO levels.

In their extensive examination of FeNO, Olin et al. [20] conducted a study involving a randomly selected cohort of healthy individuals devoid of atopy and with no history of smoking. They identified age and height as significant factors predicting FeNO levels. Similarly, Brody et al. [16] observed a linear increase in FeNO levels with age in their own random sample. Furthermore, in a multiple regression model applied to an adult reference population characterized by the absence of respiratory conditions and symptoms, as well as lifetime non-smoking status, the authors found that age and height remained independent predictors of FeNO. Conversely, in our investigation, we utilized a multiple regression model and found age, rather than height, to be a significant predictor of FeNO in the random sample. Nevertheless, akin to previous findings, both age and height emerged as significant predictors of FeNO in a healthy population excluding individuals with asthma, COPD, and allergies.

In our regression analysis, we identified age, asthma, clinical allergy, and bronchial reversibility as consistent predictors of FeNO within the randomized sample. Travers et al. [19], in their analysis, utilizing a multivariate model, delineated sex, smoking habits, height, asthma, atopy, and allergic rhinitis as significant determinants of FeNO levels. While our findings align with the conclusions drawn by Travers et al., disparities arise in the variables integrated into their models, potential biases in sample recruitment, and variations in participant age ranges. These factors may have influenced the observed significance of gender, smoking habits, and height in their study outcomes.

One of the strengths of our study lies in its comprehensive approach to data collection, which involved a wide array of clinical assessments and measurements. By conducting thorough clinical examinations and employing standardized protocols in assessing asthma and allergy parameters, we ensured the reliability and accuracy of our data. Furthermore, our sample size was large, based on a representative cohort of individuals from the general population, thereby enhancing the generalizability of our findings. Moreover, our study addressed the limitations of previous research by considering a broader range of demographic and clinical parameters, including smoking status, allergic sensitization, and respiratory conditions such as asthma and COPD. By examining the influence of these factors on FeNO levels, we provided valuable new insights into the determinants of FeNO variability within the general population.

Despite its strengths, our study has several limitations. Firstly, the cross-sectional data of this study limits our ability to establish causality between the identified factors and FeNO levels. Prospective studies would be beneficial in elucidating the temporal relationships between these variables. Secondly, while our sample size was appropriate for establishing reference values and exploring the associations between FeNO and various factors, it may have been inadequate for conducting subgroup analyses or detecting smaller effect sizes. Future studies with larger sample sizes could provide further insights into the nuances of FeNO variability within specific subgroups.

Different guidelines suggest using cut-off values rather than reference values for interpreting FeNO levels. These cut-offs vary, from 20 to 50 ppb depending on the clinical question, to indicate high FeNO levels and a possible T2 immune response [22–24]. These cut-off points are not based on reference values from the general population [24]. Our findings indicate that some of these clinically used cut-offs actually fall within the reference values for the general population. This suggests that these cut-offs are more about classification or treatment response rather than being purely abnormal. Additionally, our results demonstrate that some of these cut-offs may align with reference values from the general population, suggesting the utility of them in judging abnormality.

In conclusion, the 95^th^ percentile of FeNO ranges from 34 to 52 ppb overall, and from 26 to 37 ppb in those without clinical allergy, asthma, or COPD, depending on age. Factors such as sex, smoking, clinical allergy, atopy, asthma, and hypertension affect FeNO levels. Age, asthma, clinical allergy, and reversibility are significant predictors of FeNO in the general population. These findings are useful for interpreting FeNO measurements, considering various factors and conditions. They also provide a guide for interpreting FeNO in asthma and COPD clinics.

## Data Availability

All data produced in the present study are available upon reasonable request to the authors

## Abbreviations

BMI: body mass index
COPD: chronic obstructive pulmonary disease
FeNO: fractional exhaled nitric oxide
FEV_1_: forced expiratory volume in one second
FVC: forced vital capacity
GINA: Global Initiative for Asthma
GOLD: Global Strategy for the Diagnosis, Management and Prevention of Chronic Obstructive Lung Disease
ICS: inhaled corticosteroids
ppb: parts per billion
sIgE: specific immunoglobulin E
SPT: skin prick test
WSAS: West Sweden Asthma Study

## Notes

**Support statement** The study is supported by the VBG Group Herman Krefting Foundation for Asthma and Allergy Research (Trollhättan, Sweden), Swedish Research Council, the Swedish Heart-Lung Foundation (Stockholm, Sweden), the Swedish Asthma and Allergy Foundation (Stockholm, Sweden), Tampere Tuberculosis Foundation (Tampere, Finland), and ALF agreement (grant from the Swedish state under the agreement between the Swedish Government and the county councils, Sweden).

**Conflict of interest LL** reports personal fees from ALK, AstraZeneca, Berlin Chemie, Boehringer- Ingelheim, Chiesi, GSK, Novartis, Orion Pharma and Sanofi outside the current work. **SSÖE** reports conference-attendance related costs from Thermo Fisher Scientific outside the current work. **TP** reports fees for lectures and/or consulting from AstraZeneca, Chiesi, GSK, Novartis and Sanofi outside the current work. **HB** reports personal fees for lectures form AstraZeneca, Boehringer Ingelheim and GSK outside the current work. **NIB** reports personal fees for lectures and consulting from DBV Technologies and AstraZeneca outside the current work. **HK** reports fees for lectures and/or consulting from AstraZeneca, Boehringer-Ingelheim, Chiesi, Covis Pharma, GSK, MedScape, MSD, Novartis, Orion Pharma and Sanofi outside the current work. The rest of the authors have no conflict of interest to declare.

### Competing Interest Statement

LL reports personal fees from ALK, AstraZeneca, Berlin Chemie, Boehringer-Ingelheim, Chiesi, GSK, Novartis, Orion Pharma and Sanofi outside the current work. SSOE reports conference-attendance related costs from Thermo Fisher Scientific outside the current work. TP reports fees for lectures and/or consulting from AstraZeneca, Chiesi, GSK, Novartis and Sanofi outside the current work. HB reports personal fees for lectures form AstraZeneca, Boehringer Ingelheim and GSK outside the current work. NIB reports personal fees for lectures and consulting from DBV Technologies and AstraZeneca outside the current work. HK reports fees for lectures and/or consulting from AstraZeneca, Boehringer-Ingelheim, Chiesi, Covis Pharma, GSK, MedScape, MSD, Novartis, Orion Pharma and Sanofi outside the current work. The rest of the authors have no conflict of interest to declare.

### Funding Statement

The study was funded by the VBG Group Herman Krefting Foundation for Asthma and Allergy Research (Trollhattan, Sweden), Swedish Research Council, the Swedish Heart-Lung Foundation (Stockholm, Sweden), the Swedish Asthma and Allergy Foundation (Stockholm, Sweden), Tampere Tuberculosis Foundation (Tampere, Finland), and ALF agreement (grant from the Swedish state under the agreement between the Swedish Government and the county councils, Sweden).

### Author Declarations

All participants signed informed consent, and the study was approved by the regional ethics board in Gothenburg, Sweden which as a part of the national Swedish Ethical Review Authority that examines applications for ethics review of research involving humans and human biological material. More info can be found in: https://etikprovningsmyndigheten.se/en/

