## Supplementary file for "Reference values and determinants of fractional exhaled nitric oxide in a representative adult population in Western Sweden"

Supplementary data:

**Methods**
**Assessment of sensitization and clinical allergy**

Sensitization was assessed through the determination of sIgE levels and/or skin prick tests for 11 aeroallergens. In summary, blood samples were procured during clinical visits and subsequently preserved at -80°C. An evaluation of IgE levels against a composite of aeroallergens (Phadiatop) was then undertaken. Individuals exhibiting titers of ≥0.35 kUA/L underwent supplementary measurements for IgE antibody levels against specific allergens within the composite mixture, including cat, dog, horse, house dust mite (*Dermatophagoides pteronyssinus*, *Dermatophagoides farinae*), mold (*Cladosporium herbarum*), birch, timothy grass, and mugwort. Quantification of IgE levels was executed using the ImmunoCAP™ system (Phadia AB, Uppsala, Sweden), where IgE values equal to or surpassing 0.35 kUA/L for an individual allergen were regarded as positive. The SPTs comprised a standard panel of 11 aeroallergens (ALK, Hørsholm, Denmark), administered after a minimum antihistamine withdrawal period of 72 hours. A positive result was defined as a mean wheal diameter ≥3 mm after 15 min. *Clinical allergy* was defined by the presence of allergic sensitization (positive SPT or sIgE to any allergen; atopy), and self-reported allergic symptoms attributable to the same allergen family. These symptoms, evaluated during the clinical interview prior to the sensitization test results, included ocular manifestations, nasal discomfort, various forms of allergic nasal expressions, pruritus in the oral or pharyngeal region, respiratory challenges, exacerbation of asthma symptoms, pruritic skin rash, and disruptions in gastrointestinal function.

**Measurement of lung function and FeNO**

Spirometry measurements were conducted using a MasterScope spirometer (Jaeger, Höchberg, Germany), adhering to the guidelines stipulated by the ERS/ATS [29,30]. The Global Lung function Initiative's normal equation for Caucasians was utilized for reference values [31], enabling the calculation of Forced Expiratory Volume in 1 second (FEV_1_) percentage of predicted normal value (FEV_1_% predicted). Reversibility testing was performed 15 minutes thereafter. Methacholine challenge was executed by Spira equipment (Spira Respiratory Care Center Ltd, Hämeenlinna, Finland) adhering to ERS guidelines [32].

**Definitions of diseases**

*Asthma* was defined as meeting any of the following criteria: 1) documented diagnosis of asthma by a healthcare professional or a history of asthma accompanied by respiratory symptoms or use of asthma medication in the past 12 months; 2) respiratory symptoms or use of asthma medication in the past 12 months, coupled with a positive reversibility test (12% and 200 ml increase in FEV_1_); or 3) respiratory symptoms or use of asthma medication in the past 12 months, accompanied by a positive methacholine challenge indicative of asthma. *COPD* was defined by a post-bronchodilator FEV_1_/Forced Vital Capacity (FEV_1_/FVC) ratio of less than 0.7, along with a smoking history of 10 or more pack-years [33]. *Metabolic disease* was defined as having obesity (body mass index [BMI] ≥30 kg/m2) or any of the following self-reported conditions: hypertension, hyperlipidaemia, or diabetes [34].

**Results:**

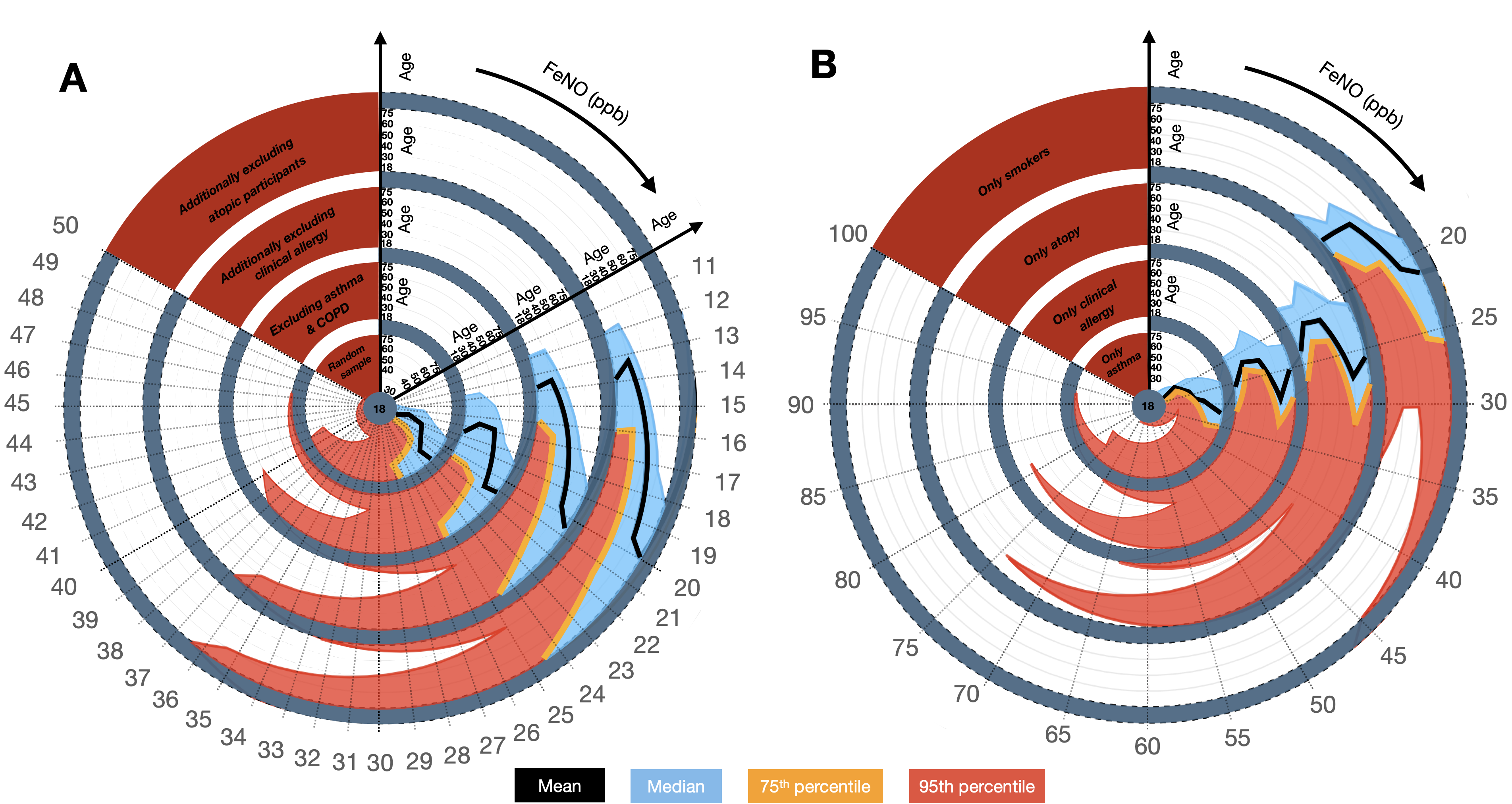

**Figure S1:** illustrates the characteristics of fractional exhaled nitric oxide (FeNO) across different age groups, delineated for: (A) the entire random sample (*N*=943), random sample participants excluding asthma and COPD patients (*n*=773), individuals additionally devoid of clinical allergy (*n*=587) , and individuals additionally devoid of atopy (*n=*519). (B) individuals with asthma (*n*=153), individuals with clinical allergy (*n*=276), atopic participants (*n*=349), and all smokers only (*n*=456). The depicted blue region demarcates the space between the median and 75^th^ percentile curves, while the red region signifies the interval between the 75^th^ percentile and 95^th^ percentile curves.

**Table S1: Factors associated with increased FeNO levels in multiple linear regression analysis in participants devoid of clinical allergy, asthma and COPD.**

|  | Coefficient B | Std. Error | Lower 95% CI | Upper 95% CI | *p-* value |
| --- | --- | --- | --- | --- | --- |
| **Demographics** |  |  |  |  |  |
| Age | 0.459 | 0.017 | 0.426 | 0.492 | **<0.001** |
| Gender | -1.745 | 0.155 | -2.049 | -1.441 | **<0.001** |
| Height | 0.330 | 0.024 | 0.284 | 0.377 | **<0.001** |
| Weight | -0.295 | 0.022 | -0.338 | -0.251 | **<0.001** |
| BMI | 0.945 | 0.066 | 0.816 | 1.074 | **<0.001** |
| Ever smoking | -0.045 | 0.068 | -0.179 | 0.089 | 0.511 |
| Current smoking | -3.543 | 0.110 | -3.759 | -3.328 | **<0.001** |
| **Comorbidities** |  |  |  |  |  |
| Hypertension | 1.094 | 0.081 | 0.936 | 1.253 | **<0.001** |
| Diabetes | -3.979 | 0.205 | -4.382 | -3.577 | **<0.001** |
| **Lung function*** |  |  |  |  |  |
| FEV_1_ pred % | -0.647 | 0.081 | -0.805 | -0.489 | **<0.001** |
| FVC | -3.089 | 1.096 | -5.238 | -0.941 | **0.005** |
| FVC pred % | 0.845 | 0.069 | 0.710 | 0.981 | **<0.001** |
| FEV_1_/FVC | 77.499 | 7.806 | 62.198 | 92.799 | **<0.001** |
| Post FEV_1_ | -4.118 | 0.631 | -5.355 | -2.880 | **<0.001** |
| Post FEV_1_ pred % | 0.556 | 0.086 | 0.387 | 0.725 | **<0.001** |
| Post FVC | 9.770 | 1.057 | 7.698 | 11.841 | **<0.001** |
| Post FVC pred % | -0.854 | 0.065 | -0.982 | -0.726 | **<0.001** |
| Post FEV_1_/FVC | -38.490 | 7.748 | -53.677 | -23.303 | **<0.001** |
| Post-pre FEV_1_ | -14.829 | 1.509 | -17.786 | -11.872 | **<0.001** |
| Reversibility % | 0.364 | 0.051 | 0.264 | 0.463 | **<0.001** |
| **R**: 0.447, R Square: 0.200, Adjusted R Square: 0.200 | | | | | |

Variables linked with the elevation of FeNO were examined through linear regression analyses encompassing a cohort devoted from asthma, COPD and allergies (n = 519). FEV1: Forced expiratory volume in 1 second, FEV1%: Percentage of predicted normal value, FVC: Forced vital capacity. *Pre FEV_1_, pre FEV_1_/FVC, and Post FEV_1_/FVC predicted % variables were excluded from the regression analysis due to multicollinearity reasons. Data are presented as the unstandardized b coefficient, standard error of b, and 95% CI.
